# Explaining national differences in the mortality of Covid-19: individual patient simulation model to investigate the effects of testing policy and other factors on apparent mortality

**DOI:** 10.1101/2020.04.02.20050633

**Authors:** Jonathan A Michaels, Matt D Stevenson

## Abstract

There has been extensive speculation on the apparent differences in mortality between countries reporting on the confirmed cases and deaths due to Covid-19. A number of explanations have been suggested, but there is no clear evidence about how apparent fatality rates may be expected to vary with the different testing regimes, admission policies and other variables. An individual patient simulation model was developed to address this question. Parameters and sensitivity analysis based upon recent international data sources for Covid-19 and results were averaged over 100 iterations for a simulated cohort of over 500,000 patients.

Different testing regimes for Covid-19 were considered; testing admitted patients only, various rates of community testing of symptomatic cases and active contact-tracing and screening.

In the base case analysis, apparent mortality ranged from 10.5% under a policy of testing only admitted patients to 0.4% with intensive contact tracing and community testing. These findings were sensitive to assumptions regarding admission rates and the rate of spread, with more selective admission policies and suppression of spread increasing the apparent mortality and the potential for apparent mortality rates to exceed 18% under some circumstances. Under all scenarios the proportion of patients tested in the community had the greatest impact on apparent mortality.

Whilst differences in mortality due to health service and demographic factors cannot be excluded, the current international differences in reported mortality are all consistent with differences in practice regarding screening, community testing and admission policies.

## INTRODUCTION

The current Covid-19 pandemic has been the subject of more open and rapid availability of data than any previous disease. This has, inevitably, led to international comparisons of the spread and outcome of the disease, with widespread media speculation regarding the apparent differences in mortality between similar European countries. On the face of it, the differences are stark. On 2^nd^ April 2020, Germany reported 85,063 confirmed cases with 1,111 deaths (1.3%), whereas three days earlier the comparable figures from Spain had shown 7,716 deaths from 87,956 confirmed cases (8.8%) and three days before that Italy had reported 9,134 deaths from 86,498 cases (10.1%).[1] While these may relate to differences in demographics, treatment or healthcare policy, the apparent mortality may also be altered by differences in testing policy and in changes in the rate of spread brought about by social distancing policies.

In estimating mortality in the early stages of a rapidly spreading infection there are important potential confounding factors. Early under-ascertainment with a failure to identify mild, moderate or asymptomatic cases in the community, may lead to an overestimate of mortality. Conversely, the rapid rise in identified cases with unknown outcomes may lead to an underestimate as confirmed cases are identified a considerable time before deaths occur (right-censoring). This paper describes a simulation model of the effect of different testing regimes on the apparent mortality in the early stages of exponential spread of a pandemic, and compares this to reported international variation in mortality rates for Covid-19.

## METHOD

An individual patient level simulation model was developed in R (R Foundation for Statistical Computing, Vienna) to consider the effects of different testing policies, spread parameters and hospital admission rates on the apparent mortality of a pandemic in the early phase of exponential spread. A series of testing scenarios were considered;

- Testing restricted to those admitted to hospital with severe disease.
- The addition of community testing for virus in a proportion of symptomatic patients. Scenarios considered testing rates of 10%, 25%, 50%, 75% and 90%.
- Active tracing of known contacts and testing to identify symptomatic and asymptomatic infections.

The first 19 doubling cycles of spread were considered, resulting in over 500,000 simulated cases. Apparent mortality for each testing scenario was estimated, based upon averaging 100 iterations to allow for Monte Carlo error, using a common seed to provide consistency across model runs. Model estimates under the different scenarios were compared to the apparent mortality rates based upon published national data, for those countries with the greatest number of reported cases of Covid-19. The structure of the model is shown diagrammatically in Figure 1.

**Figure 1.**
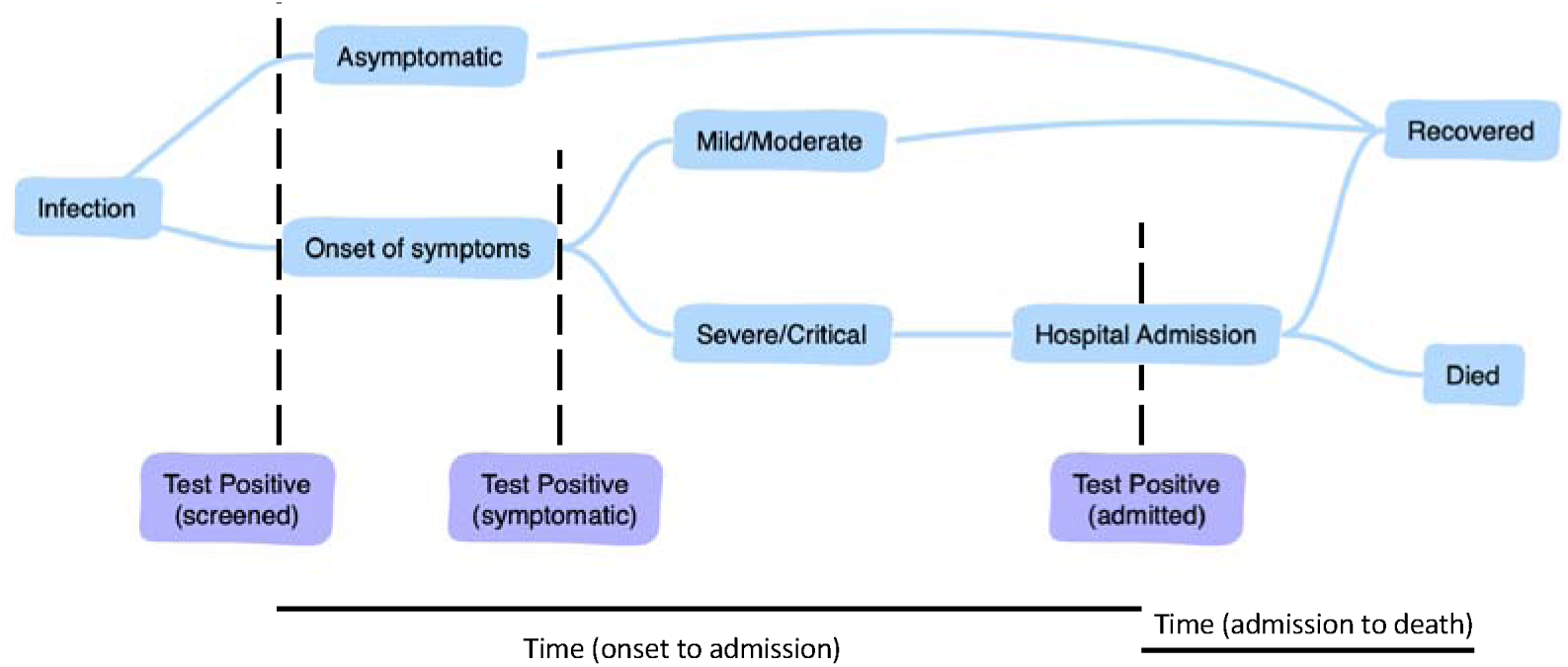
Structure of the simulation model for Covid-19.

### Parameters and assumptions

A summary of the base case estimates for the required parameters, ranges used in the sensitivity analyses, and sources of data are provided in Table 1.

**Table 1.** Parameter values used in the base case analysis and sensitivity analysis.

| Parameter | Base case | Sensitivity analysis | Source |
| --- | --- | --- | --- |
| Underlying IFR | 0.66% | 0.2% to 5% | [1, 2, 4, 5] |
| Proportion that are severe/critical requiring admission | 9.2% | 4.6% to 18.4% | [2, 7] |
| Days (IQR) onset to admission | 5.3 | 1.9 to 6.8 | [4] |
| Days (IQR) admission to death | 9.1 | 6.7 to 13.7 | [4] |
| Asymptomatic proportion | 17.9% | 7.7%-53.5% | [9, 10] |
| Doubling time (days) | 6 | 2 to 10 | [1] |
| Testing policy | 50% of community cases | 0 to 90% of community cases and contact tracing | Scenario analysis |
| Infections identifiable through contact tracing | 50% | 20% to 80% | Assumption |

#### Infection fatality rate (IFR)

There is considerable doubt about the underlying IFR of Covid-19. The apparent case fatality rate based upon reported deaths[1] is around 4.8% and an estimate based upon ‘completed’ cases from China suggests a figure of 2.3%.[2] Assumptions regarding asymptomatic or untested cases result in very much lower estimates for IFR that have been used in previous models.[3] For the base case in this study a published estimate of 0.66%[4] was used, with sensitivity analysis covering a wide range from 0.2% to 4.8%.[5]

#### Rate of spread

The rate of spread was assumed to be exponential in the early stages and expressed in terms of days required for numbers to double, with a base case estimate of six days and range from 2 to 10 days.[1]

#### Rate of hospital admission

The rate of hospital admission was modelled as a multiple of the number of deaths. This was chosen as it is a figure that can be rapidly and more accurately ascertained from local data, whereas the actual admission rate as a proportion of infected cases is uncertain due to unknown incidence in the community. In the early data from China the overall admission rate was approximately eight patients for every death.[6] However, these data include a significant proportion of patients with mild/moderate disease, who may have been admitted for isolation. Estimates based upon only severe and critical cases put the ratio at nearer to 4.[7]

#### Delay from onset to admission and admission to fatality

The distribution of time intervals between onset of symptoms and hospital admission and between admission and death were obtained from models based upon the Chinese data.[4] Separate estimates were provided for survivors and non-survivors; however, it seems likely that the survivor data is distorted by those with less severe disease, who may have been admitted early for isolation.[8] Thus, for the base case, the time from onset to admission for all cases was based upon the non-survivor data. Sensitivity analysis included the full range reported for both groups. The time from admission to death was based upon the modelled distributions from same data.[4]

#### Proportion of asymptomatic cases

The proportion of infections that remain asymptomatic is unknown, but has been estimated at 17.9% from the screened populations aboard the Diamond Princess cruise ship[9] and at 33%, based on Japanese nationals evacuated from Wuhan.[10]

### Testing regimes

It is assumed that all hospitalised patients will be tested. Various scenarios have been considered with 10% to 90% of symptomatic cases in the community being tested. In addition, a policy of tracing and testing known contacts has been considered. In this scenario it is assumed that asymptomatic cases would not otherwise have been identified and symptomatic cases will be identified two days earlier than would otherwise have been the case. The proportion of infections that are amenable to identification through contact tracing is dependent upon the local circumstances. Under strict lock-down it might be expected that most contacts would be identifiable, whereas with free movement on public transport this may be impossible. Thus, an assumption of 50% was made with minimum and maximum values of 20% and 80%.

## RESULTS

Under the base case assumptions, the apparent mortality estimated from confirmed cases and known deaths is 5.3% with the assumption of 50% community testing of symptomatic patients, and ranges from 10.5%, if testing is restricted to hospitalised patients, down to 0.4% with intensive contact tracing and community testing (Table 2). Figure 2 illustrates the one-way sensitivity analysis of the key parameters as specified in Table 1 (see Appendix 1 for the full results of sensitivity analysis). The rate of community testing and the ratio of hospitalised patients have the greatest effect and are considered in a two-way sensitivity analysis (see Table 2).

**Table 2.**
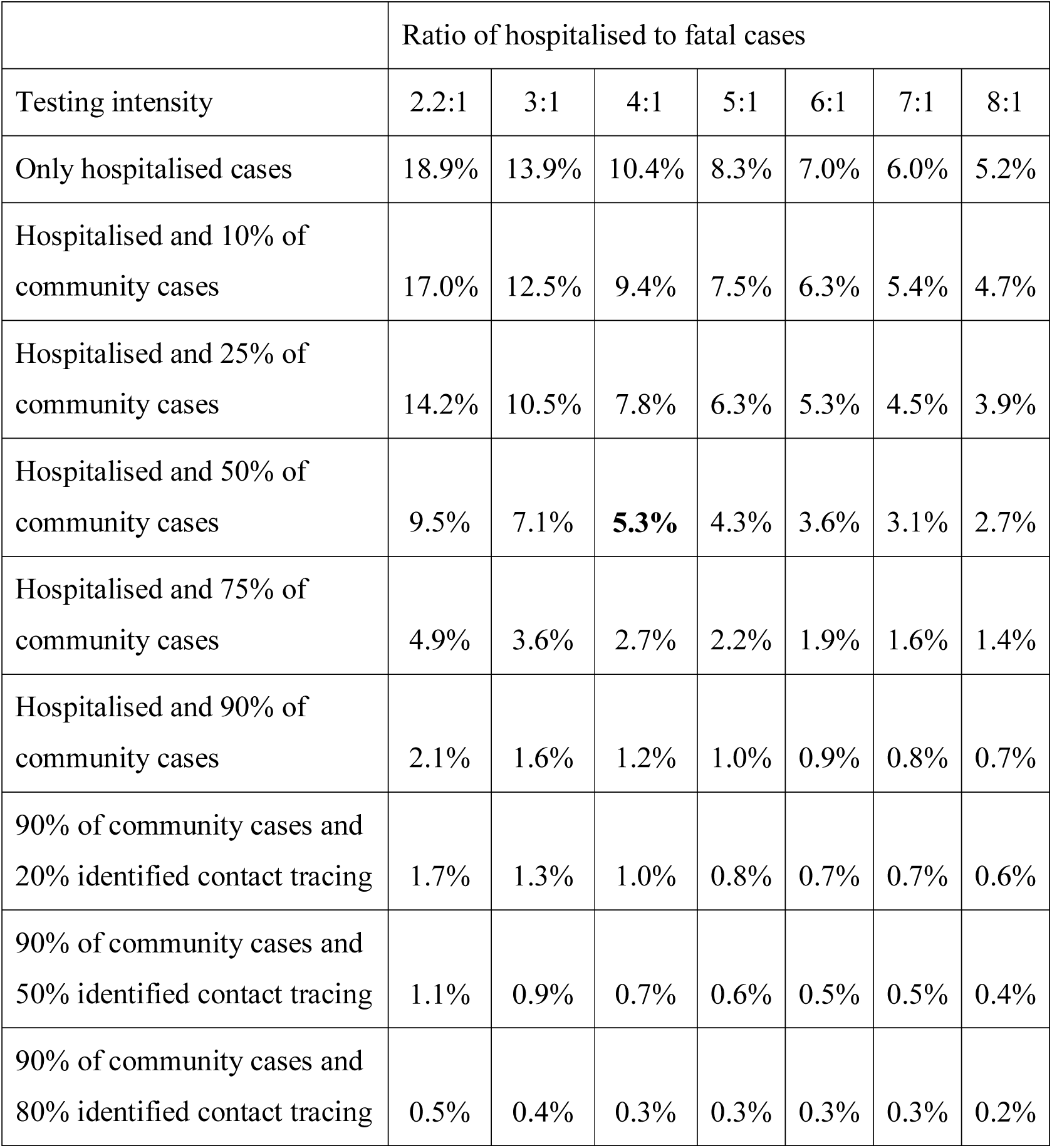
Two-way sensitivity analysis of testing intensity and ratio of hospitalised to fatal cases (base case in bold) in relation to apparent mortality.

**Figure 2.**
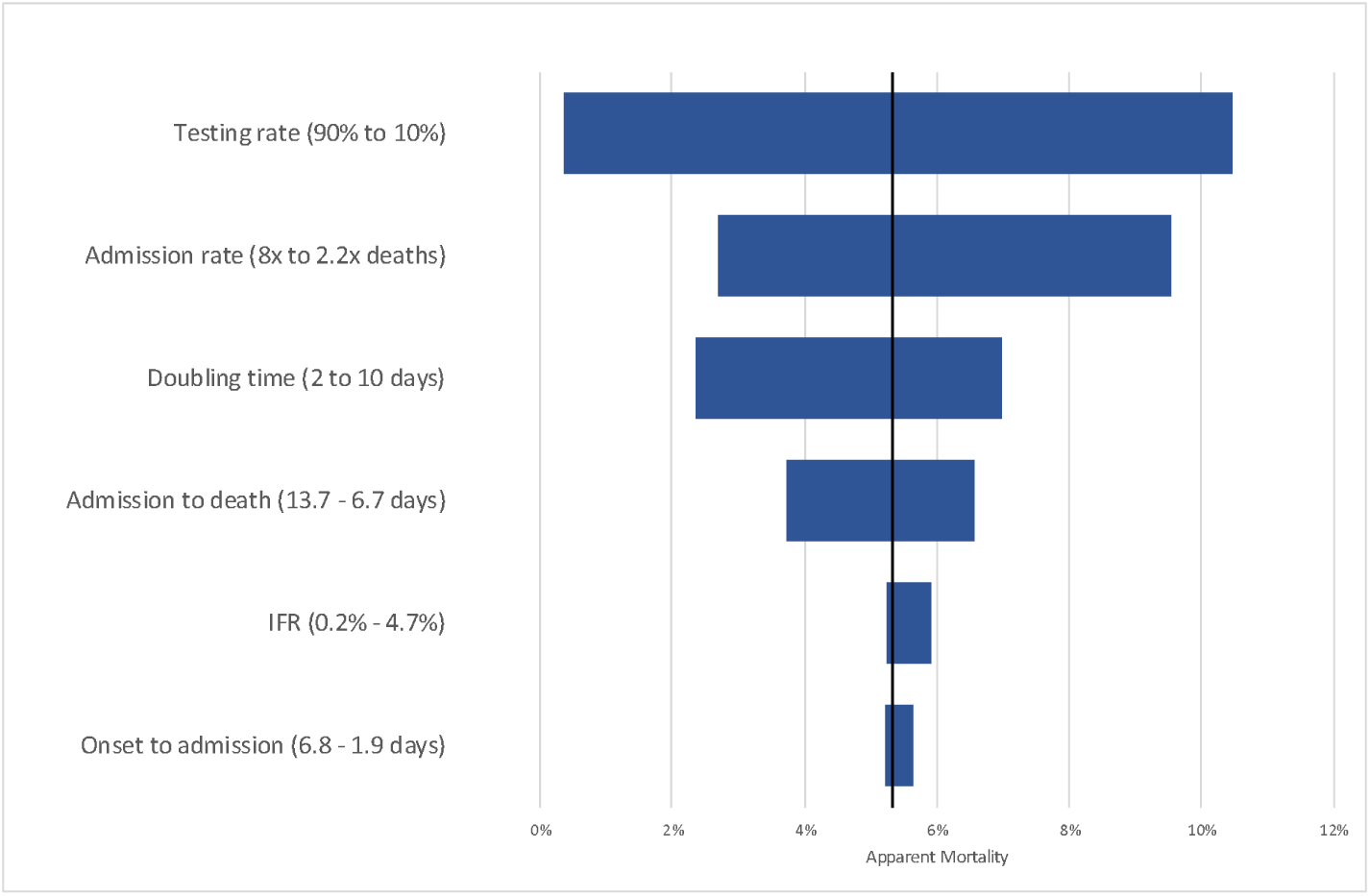
Tornado diagram of the results of a one-way sensitivity analysis based upon the parameter ranges in Table 1. The vertical line represents the base case (5.3%).

The apparent fatality rates from Covid-19 in those countries with the greatest number of cases is shown in Figure 3, with the modelled estimates different community testing regimes.

**Figure 3.**
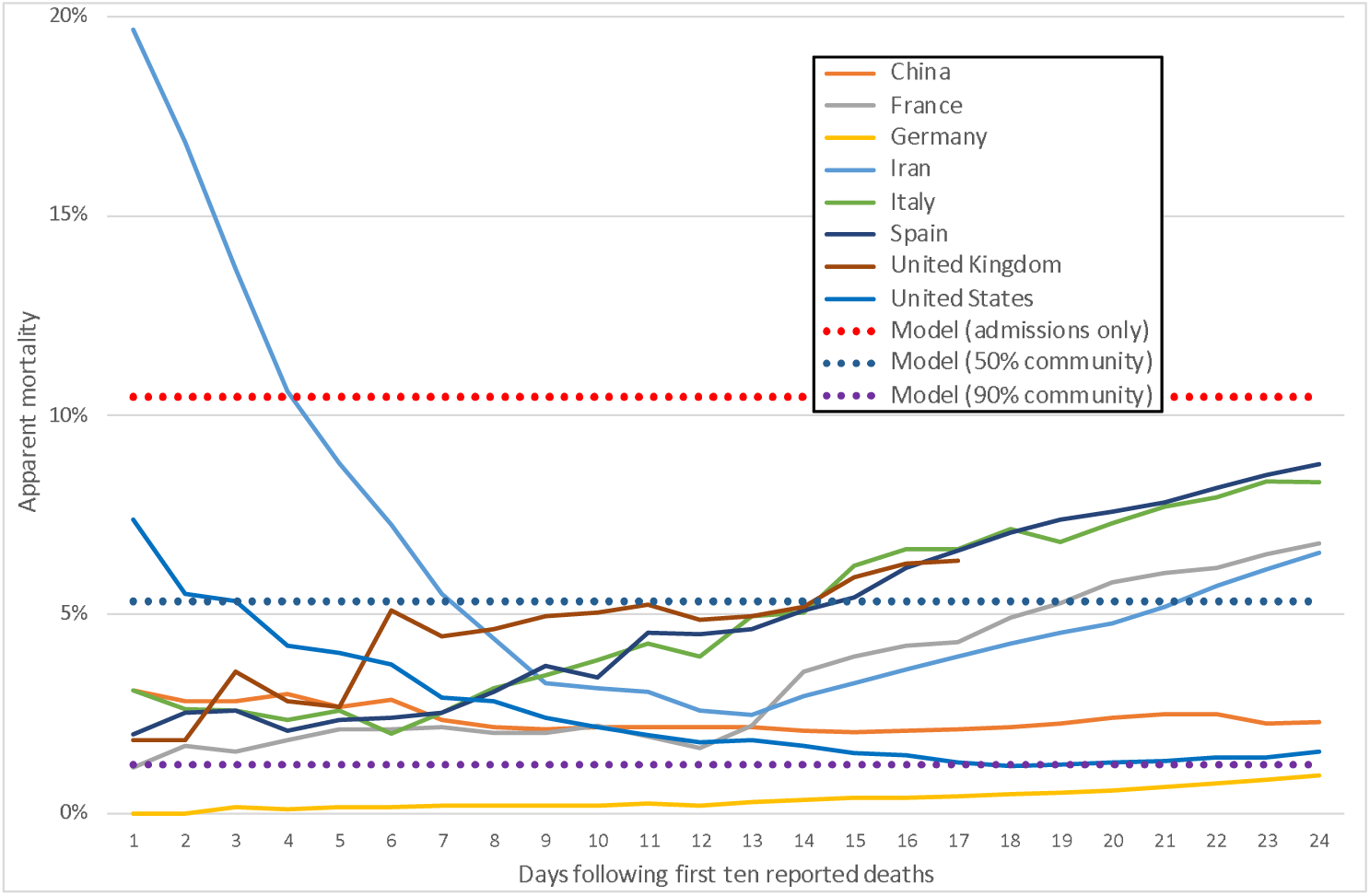
Apparent fatality rate from Covid-19 for 24 days after the first 10 deaths were reported in those countries with the highest reported rates, compared to modelled apparent mortality with differing rates of community testing.

## DISCUSSION

The modelling demonstrates that the large differences between countries in apparent Covid-19 mortality rates are compatible with the effects that might be expected with different testing policy in the early stages of a pandemic. While it is not possible to rule out differences in outcome due to demographic factors or aspects of the provision of health services, care is needed in drawing conclusions about such differences. The effects of more stringent testing regimes and tighter hospital admission policies are likely to exaggerate apparent fatality rates. High pressure on services, due to rapidly increasing demands, may affect both of these through limitations in staff, equipment and test kits for community testing as well as increasing the threshold of severity for hospital admission. Paradoxically, since the effect of incomplete case ascertainment is partly counter-balanced by the effect of right-censoring, successful attempts as suppression, which reduce the impact of right-censoring, may appear to exaggerate estimates of mortality.

It has been suggested that the use of historical numbers of confirmed cases, 14 days prior to fatality rates, as the denominator may provide more accurate estimates than basing rates on the most recent deaths and confirmed cases.[11] Due to the skewed distribution of survival a more sophisticated estimate may be obtained by using weighted averages over a longer period. However, neither of these would account for the other factors described, such as under-ascertainment, that might distort estimates in the opposite direction.

Sensitivity analysis suggests that the underlying IFR has relatively little effect on apparent mortality, since the uncertainty largely relates to the number of asymptomatic or mild cases that remain unidentified. Conversely, this implies that the IFR will remain uncertain, as demonstrated by the widely differing estimates,[4] until the result of more extensive population testing become available. Another implication is that it is likely that those countries reporting higher mortality rates have a large number of unidentified cases and there is an urgent need to improve our understanding of this in order to predict future trends and the effects of suppression measures. If the apparent differences between German and Italian mortality were to be entirely related to differences in admission and testing criteria, then the implication is that there were at least one million unconfirmed infections in Italy by 30 March 2020.

As with any modelling, this study is limited by the available information, but the general findings remain robust across a range of sensitivity analyses. Some of the parameters and assumptions are based upon very limited data in selected populations, such as specific nations and cruise ship passengers, and may not be representative of the wider populations. There may also be some concerns about the accuracy of reported data and, in particular, the reported deaths are largely based upon hospital admissions, whereas there may be a significant mortality amongst untested people in care homes and other settings that are excluded from the data.

In conclusion, this study demonstrates the potential dangers of speculation and over-interpretation of apparent differences in mortality, without adjustment for differences in testing policy, admission rates and rate of spread. It also provides a basis for understanding the likely effects of these differences in practice on reported mortality rates.

## Data Availability

Model code will be available on request from the author.

## Appendix 1: Results of sensitivity analysis

| Case | DT | IFR | As | AR | TTA | TTD | S1 | S2 | S3 | S4 | S5 | S6 | S7 | S8 | S9 |
| --- | --- | --- | --- | --- | --- | --- | --- | --- | --- | --- | --- | --- | --- | --- | --- |
| Doubling time 2 days | 2 | 0.0066 | 0.179 | 4 | 5.3 | 9.1 | 4.7% | 4.2% | 3.5% | 2.4% | 1.2% | 0.5% | 0.4% | 0.3% | 0.1% |
| Doubling time 3 days | 3 | 0.0066 | 0.179 | 4 | 5.3 | 9.1 | 6.4% | 5.7% | 4.8% | 3.2% | 1.7% | 0.7% | 0.6% | 0.4% | 0.2% |
| Doubling time 4 days | 4 | 0.0066 | 0.179 | 4 | 5.3 | 9.1 | 7.9% | 7.1% | 5.9% | 4.0% | 2.1% | 0.9% | 0.7% | 0.5% | 0.2% |
| Doubling time 5 days | 5 | 0.0066 | 0.179 | 4 | 5.3 | 9.1 | 9.2% | 8.3% | 7.0% | 4.7% | 2.4% | 1.1% | 0.9% | 0.6% | 0.3% |
| Doubling time 6 days (base case) | 6 | 0.0066 | 0.179 | 4 | 5.3 | 9.1 | 10.5% | 9.4% | 7.9% | 5.3% | 2.8% | 1.2% | 1.0% | 0.7% | 0.4% |
| Doubling time 7 days | 7 | 0.0066 | 0.179 | 4 | 5.3 | 9.1 | 11.4% | 10.3% | 8.6% | 5.8% | 3.0% | 1.4% | 1.1% | 0.8% | 0.4% |
| Doubling time 8 days | 8 | 0.0066 | 0.179 | 4 | 5.3 | 9.1 | 12.3% | 11.1% | 9.3% | 6.3% | 3.3% | 1.5% | 1.2% | 0.8% | 0.4% |
| Doubling time 9 days | 9 | 0.0066 | 0.179 | 4 | 5.3 | 9.1 | 13.1% | 11.8% | 9.9% | 6.7% | 3.5% | 1.6% | 1.3% | 0.9% | 0.5% |
| Doubling time 10 days | 10 | 0.0066 | 0.179 | 4 | 5.3 | 9.1 | 13.7% | 12.3% | 10.3% | 7.0% | 3.7% | 1.6% | 1.4% | 0.9% | 0.5% |
| Low asymptomatic rate | 6 | 0.0066 | 0.077 | 4 | 5.3 | 9.1 | 10.5% | 9.4% | 7.9% | 5.3% | 2.8% | 1.2% | 1.0% | 0.7% | 0.3% |
| Higher asymptomatic rate | 6 | 0.0066 | 0.535 | 4 | 5.3 | 9.1 | 10.4% | 9.4% | 7.9% | 5.4% | 2.9% | 1.4% | 1.1% | 0.7% | 0.4% |
| Low admission rate (2.2) | 6 | 0.0066 | 0.179 | 2.2 | 5.3 | 9.1 | 18.9% | 17.0% | 14.2% | 9.5% | 4.9% | 2.1% | 1.7% | 1.1% | 0.5% |
| Admission rate 3 | 6 | 0.0066 | 0.179 | 3 | 5.3 | 9.1 | 13.9% | 12.5% | 10.5% | 7.1% | 3.6% | 1.6% | 1.3% | 0.9% | 0.4% |
| Admission rate 5 | 6 | 0.0066 | 0.179 | 5 | 5.3 | 9.1 | 8.3% | 7.5% | 6.3% | 4.3% | 2.2% | 1.0% | 0.8% | 0.6% | 0.3% |
| Admission rate 6 | 6 | 0.0066 | 0.179 | 6 | 5.3 | 9.1 | 7.0% | 6.3% | 5.3% | 3.6% | 1.9% | 0.9% | 0.7% | 0.5% | 0.3% |
| Admission rate 7 | 6 | 0.0066 | 0.179 | 7 | 5.3 | 9.1 | 6.0% | 10.5% | 4.5% | 3.1% | 1.6% | 0.8% | 0.7% | 0.5% | 0.3% |
| Highest admission rate | 6 | 0.0066 | 0.179 | 8 | 5.3 | 9.1 | 5.2% | 4.7% | 3.9% | 2.7% | 1.4% | 0.7% | 0.6% | 0.4% | 0.2% |
| Low Time to death | 6 | 0.0066 | 0.179 | 4 | 5.3 | 6.7 | 12.9% | 11.6% | 9.7% | 6.6% | 3.4% | 1.5% | 1.2% | 0.8% | 0.4% |
| High Time to death | 6 | 0.0066 | 0.179 | 4 | 5.3 | 13.7 | 7.3% | 6.6% | 5.5% | 3.7% | 1.9% | 0.9% | 0.7% | 0.5% | 0.2% |
| Low Time to admission | 6 | 0.0066 | 0.179 | 4 | 1.9 | 9.1 | 11.0% | 9.9% | 8.3% | 5.6% | 3.0% | 1.4% | 1.1% | 0.8% | 0.4% |
| High Time to admission | 6 | 0.0066 | 0.179 | 4 | 6.8 | 9.1 | 10.3% | 9.3% | 7.8% | 5.2% | 2.7% | 1.2% | 1.0% | 0.6% | 0.3% |
| Low IFR | 6 | 0.002 | 0.179 | 4 | 5.3 | 9.1 | 10.4% | 9.4% | 7.8% | 5.3% | 2.7% | 1.1% | 0.9% | 0.6% | 0.3% |
| Intermediate IFR | 6 | 0.004 | 0.179 | 4 | 5.3 | 9.1 | 10.4% | 9.4% | 7.8% | 5.3% | 2.7% | 1.1% | 0.9% | 0.6% | 0.3% |
| High IFR | 6 | 0.023 | 0.179 | 4 | 5.3 | 9.1 | 10.4% | 9.5% | 8.0% | 5.6% | 3.1% | 1.7% | 1.4% | 1.1% | 0.7% |
| Maximum IFR | 6 | 0.048 | 0.179 | 4 | 5.3 | 9.1 | 10.4% | 9.5% | 8.2% | 5.9% | 3.7% | 2.4% | 2.1% | 1.7% | 1.2% |

### Parameters

DT Doubling time (days)

IFR Infection fatality rate

As Proportion asymptomatic

AR Admission rate (multiplier for IFR)

TTA Time from onset to admission

TTD Time from admission to death

### Scenarios

S1 Testing only admitted cases

S2 Testing all admitted cases and 10% of symptomatic cases

S3 Testing all admitted cases and 25% of symptomatic cases

S4 Testing all admitted cases and 50% of symptomatic cases

S5 Testing all admitted cases and 75% of symptomatic cases

S6 Testing all admitted cases and 90% of symptomatic cases

S7 Testing all admitted cases, 90% of symptomatic cases and 20% identification through contact tracing

S8 Testing all admitted cases, 90% of symptomatic cases and 50% identification through contact tracing

S9 Testing all admitted cases, 90% of symptomatic cases and 80% identification through contact tracing

### Base case highlighted

